# WaveGate-nnU-Net: Frequency-Domain Inclusion Preserves Segmentation Accuracy under Reduced Training Data in Nasal and Paranasal Sinus CT

**DOI:** 10.64898/2026.09.10.26362809

**Authors:** Yu-Chen Chen, Shu-Yen Wan

## Abstract

**Background and Objectives:** Automatic segmentation of the nasal cavity and paranasal sinuses from CT aids diagnosis and surgical planning, but clinical datasets in this domain remain small, which can affect training stability and evaluation validity. This study investigates whether frequency-domain representation learning can be incorporated, in a lightweight form, into the self-configuring nnU-Net framework, and whether the resulting performance improvement survives rigorous statistical validation.

**Methods:** We constructed an nnU-Net with frequency inclusion, adding a multi-scale wavelet branch and a cross-domain attention module at the network bottleneck following a previously reported frequency-spatial dual-stream formulation, and corrected a zero-initialization gating deadlock in that formulation, using the corrected model (v2) as a common base. On this base, we evaluated three variants: X1, an input-conditioned learnable spectral gate applied to 24 wavelet sub-bands; X2, which adds a boundary-distance training objective; and WaveGate-nnU-Net (hereafter WaveGate), the proposed model combining both. Experiments used the 130-volume NasalSeg CT dataset, under a full 91/19/20 protocol and a 3-fold cross-validation in which the training set for each fold was reduced by approximately 19%, from 91 to 73-74 cases.

**Results:** Under the full protocol, all variants showed a small but statistically detectable improvement over a matched nnU-Net baseline. WaveGate achieved a Dice score of 95.78% versus 95.59% for the baseline, an improvement of 0.19 percentage points (95% CI [0.09, 0.29]; Holm-adjusted p = 0.004), and WaveGate also reduced average surface distance (ASD) from 0.252 mm to 0.235 mm. WaveGate was statistically equivalent to either individual component within a margin of ±0.10 percentage points, suggesting a shared performance ceiling rather than a complementary gain. Under reduced training data, baseline Dice dropped by 2.71 percentage points (95.59% to 92.88%), while WaveGate remained essentially unchanged (95.78% to 95.75%), a cross-fold advantage of +2.88 percentage points (fold-level 95% CI [0.80, 4.96]) that held across all 20 test cases (sign test, p = 1.9×10⁻⁶) and was corroborated by average surface distance.

**Conclusions:** The practical value of this combined approach lies primarily in within-distribution data efficiency, rather than in maximizing peak segmentation accuracy.

## 1. Introduction

Chronic rhinosinusitis and related sinonasal diseases are common, and CT remains the routine modality for diagnosis, surgical planning, and postoperative follow-up. Manual delineation of the nasal-sinus complex is slow and subject to inter-observer variability, motivating automated segmentation. Datasets for this anatomy are small by deep-learning standards — the public NasalSeg dataset [1] contains 130 annotated volumes — reflecting the normal condition of clinical imaging; recent work argues that the relevant question for 3D medical segmentation is often how gracefully a method degrades under limited data, not what it achieves with abundant data [2].

Within this setting, nnU-Net [3] is a self-configuring pipeline that derives preprocessing, patch/batch geometry, augmentation, and training schedule from dataset fingerprints, and remains a strong, hard-to-beat baseline. Any proposed mechanism therefore faces a natural test: does it still help inside nnU-Net, where many configuration-related sources of performance variation are already controlled? We position this work relative to the wider architecture landscape — attention-gated [4], Transformer [5], state-space [6, 7], and promptable foundation models [8, 9] — as well as large-scale, nnU-Net-based tools trained on extensive datasets, such as TotalSegmentator [10] — rather than benchmarking directly against these, since direct comparison would confound architecture with training protocol, data volume, and preprocessing.

Frequency-domain modeling has a plausible anatomical rationale here: thin bony walls and narrow air channels manifest as high-frequency structure, while low-frequency sub-bands carry global anatomical layout [11]. One recent frequency-spatial dual-stream formulation for sinonasal CT (AFS-DSN [12]) combined a multi-scale wavelet branch (db1, db2, db4; 24 sub-bands) with a spatial U-Net encoder-decoder through cross-domain attention and an adaptive router. We take its frequency-domain components, rather than the network as a whole, as the starting point for an nnU-Net with frequency inclusion, because multi-scale wavelet decomposition matches the multi-scale character of sinonasal anatomy and is one of the few frequency-domain treatments evaluated directly on this dataset.

Accordingly, this study investigates whether frequency-domain mechanisms of this kind can be incorporated into nnU-Net in a lightweight form and whether any resulting improvement remains reproducible under statistical validation. Under full training data, the answer is a qualified yes: real but small (roughly +0.2 pp Dice), without requiring the 414.57M-parameter capacity of the full dual-stream network of [12]. Under reduced training data, the answer is considerably stronger and constitutes our main result: the augmented model is nearly insensitive to an approximately 19% reduction in training cases, while the plain nnU-Net baseline loses 2.71 pp. Earlier single-split gains did not survive multi-split validation, the failure mode Varoquaux [13] describes for small samples, motivating us to treat single-fold results as exploratory evidence only.

The contributions of this work are: (1) a Protocol/Adapter framework that brings heterogeneous architectures into one verifiable experimental protocol; (2) a primary architectural ablation (X1, learnable spectral gating) and an auxiliary objective control (X2, boundary training objective) on a common base, plus their combination, the proposed WaveGate-nnU-Net model, each mechanistically isolated; and (3) a genuine 3-fold reduced-data validation covering all 110 development cases exactly once as validation, with 73-74 training cases per fold, establishing within-distribution data efficiency as the core finding.

## 2. Methods

### 2.1 Dataset and experimental protocols

All experiments use NasalSeg [1], 130 3D nasal and paranasal sinus CT volumes with voxel-wise annotation of five structures, merged into a single foreground class for binary segmentation, matching the surgical-planning objective of delineating the overall nasal-sinus complex boundary. We define two protocols on this dataset. Protocol A (full-data) partitions the data into 91 training, 19 validation, and 20 test cases with a fixed seed, and is used for model development and full-data comparison. Protocol B (reduced-data, 3-fold) repartitions the same 110 development cases (91+19) into three folds of 37/37/36 for validation, giving training sets of 73-74 cases each (an approximately 19% reduction from Protocol A), all evaluated on the same held-out 20-case test set; this is the basis of the paper’s main result. Absolute performance differences between Protocols A and B should be interpreted in light of their different training-set sizes; Protocol B was designed specifically to assess robustness to reduced training data. Preprocessing, patch geometry, augmentation, and optimization follow nnU-Net’s self-configured plan and are held identical across all compared variants. For this dataset the self-configured plan uses a patch size of 128×128×128 with six encoder stages of 32/64/128/256/320/320 channels; the last two stages downsample in-plane only (stride [1, 2, 2]), so the bottleneck feature is anisotropic at 320×16×4×4. Evaluation uses Dice, IoU, average surface distance (ASD), and normalized surface Dice at 0.5/1.0/2.0 mm, computed with each volume’s anisotropic physical spacing.

**Figure 1.**
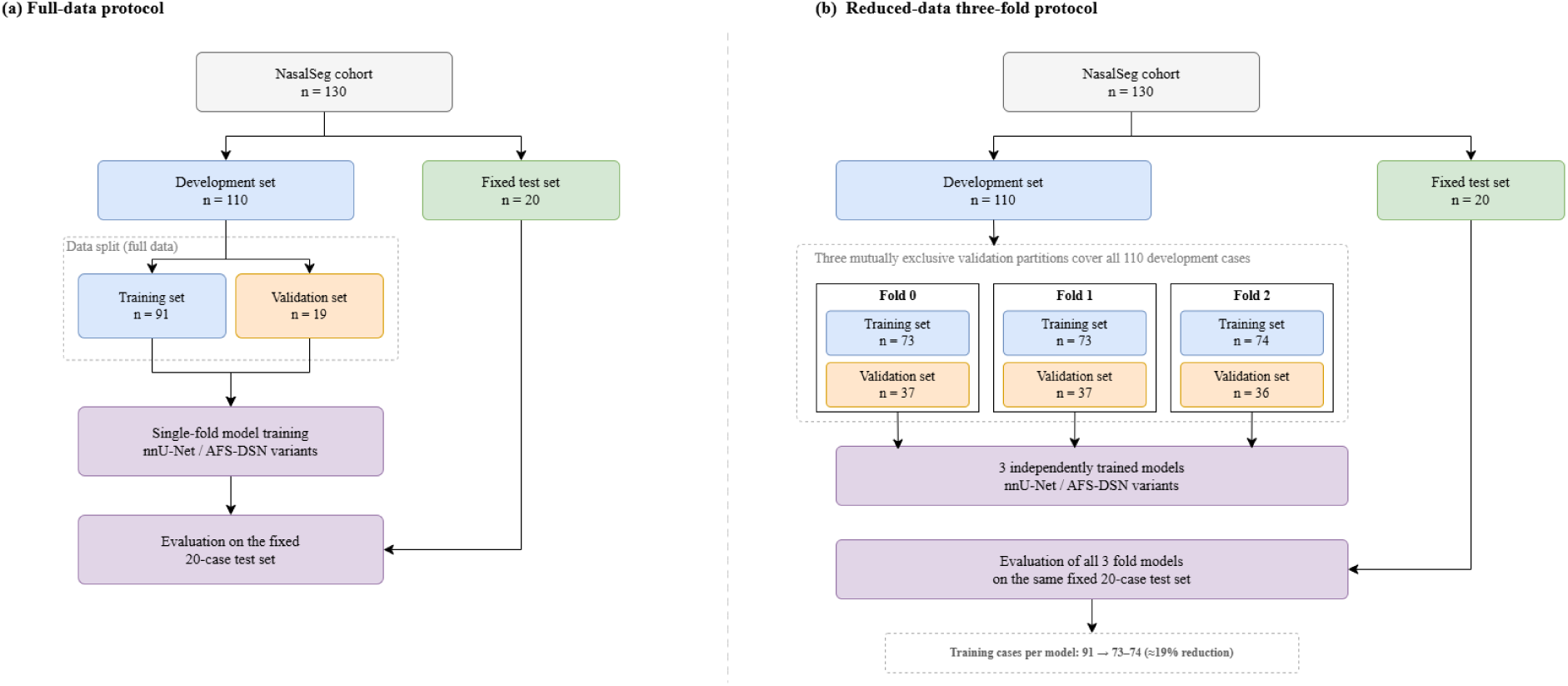
Experimental design comparison. (a) The full-data protocol: a single 91/19/20 split used for the primary screening comparison (Protocol A). (b) The reduced-data three-fold protocol: the 110-case development set is partitioned into three mutually exclusive validation folds, each defining a distinct 73–74-case training set; all three resulting models are evaluated on the same fixed 20-case test set used in panel (a).

### 2.2 Base integration and model variants

A wavelet frequency branch, cross-domain attention, and an adaptive router, following [12], were inserted into the nnU-Net encoder-decoder at the bottleneck, giving an nnU-Net with frequency inclusion. In the first integration (v1), zero-initialized residual gating coefficients produced a training deadlock (95.50% Dice, below the 95.59% nnU-Net control); re-initializing these coefficients to 0.1 (v2) restored stable training (95.71%); this corrected configuration, WaveGate-Base, serves as the common base for all subsequent variants. Only the initial value was changed: the coefficients remain learnable as in [12], and the architecture, data, splits, patch geometry, and epoch budget are identical between v1 and v2, so the comparison isolates the effect of the initialization alone.

**Figure 2.**
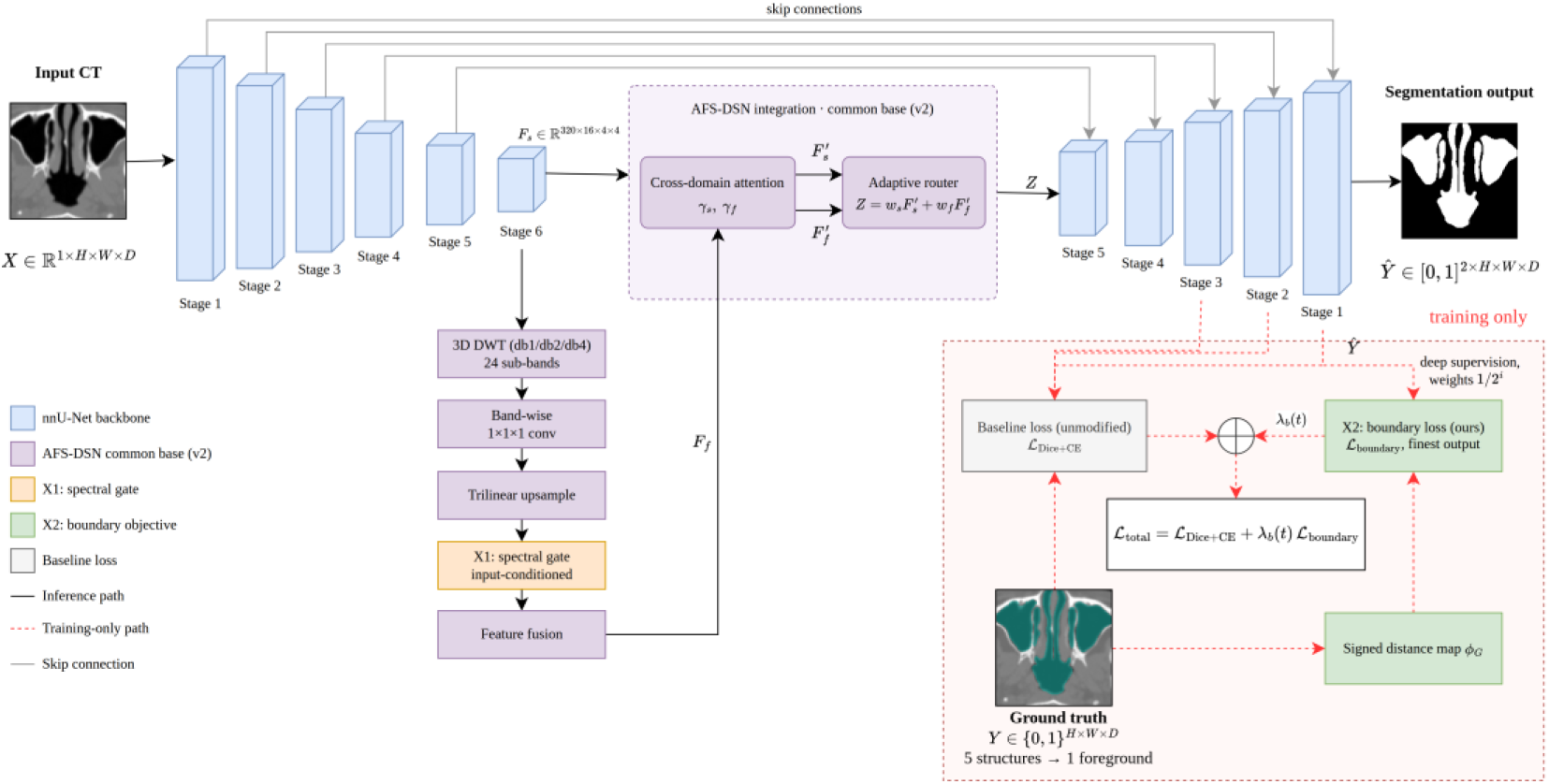
Frequency inclusion within nnU-Net. The nnU-Net backbone (blue) is unmodified: six encoder stages over a 128×128×128 patch, the last two downsampling in-plane only, giving the bottleneck feature F_s at 320×16×4×4. The frequency-inclusive common base (purple) decomposes F_s by 3D DWT into 24 wavelet sub-bands (db1, db2, db4), applies band-wise 1×1×1 convolutions with trilinear resampling, and fuses the resulting frequency representation F_f with the spatial stream through bidirectional cross-domain attention with gated residuals (γ_s, γ_f) and an adaptive router. X1 (orange) replaces the globally fixed band weights of the common base with an input-conditioned spectral gate driven by log-compressed band-energy statistics. X2 (green) adds a boundary-distance term computed from the signed distance map of the ground truth, applied at the finest decoder scale only and annealed by λ_b(t) from 0 to 0.2 over the first 60 epochs. WaveGate combines X1 and X2 and constitutes the proposed model. Solid arrows denote the inference path; dashed red arrows denote training-only computation. Three deep-supervision heads are drawn for clarity; the loss is applied at every supervised scale. Overlays show an axial slice of NasalSeg_104: ground truth in teal on the CT, network output as a binary mask.

The frequency branch decomposes the bottleneck feature into 24 sub-bands across three Daubechies bases (Eq. 1). Each sub-band is then passed through its own 1×1×1 convolution and trilinearly resampled back to the bottleneck resolution, giving the 24 band features F_k that all subsequent frequency-domain operations act on. The spatial and frequency streams are then refined by bidirectional cross-domain attention, scaled by 1/√d_k (Eq. 2), and each stream is updated by a residual connection scaled by a learnable scalar γ_s or γ_f (Eq. 3), following [12]. These are the coefficients whose zero-initialization caused the v1 deadlock: scaling the residual branch by γ makes the gradient with respect to γ itself vanish when γ starts at zero, so the frequency pathway never begins to contribute; re-initializing γ to 0.1 removes this degenerate starting point. The two refined streams are then combined by the adaptive router, which computes global statistics of both streams and applies a softmax to obtain content-dependent mixing weights (Eq. 4).

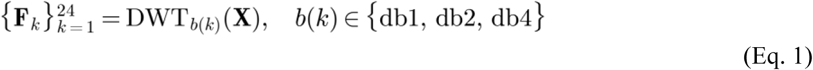

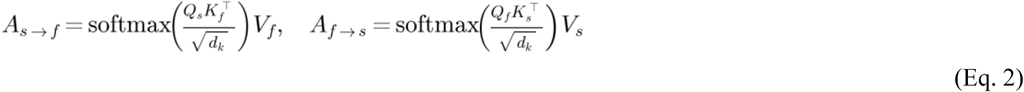

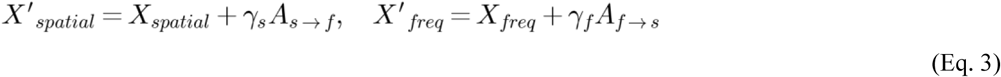

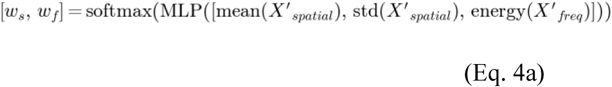

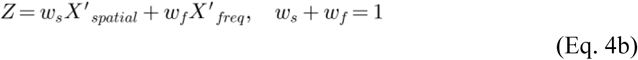

On this base, the spectral-gate variant WaveGate-X1 (abbreviated X1 below) replaces the globally fixed, learnable band weights of the WaveGate-Base configuration with an input-conditioned spectral gate: per-sub-band energy statistics are passed through a small MLP with sigmoid output to produce 24 gate values that rescale the corresponding band features before cross-domain attention and adaptive-router fusion, otherwise unchanged from v2. A synthetic differentiation test confirmed that the spectral gate was trainable by gradient descent.

**Figure 3.**
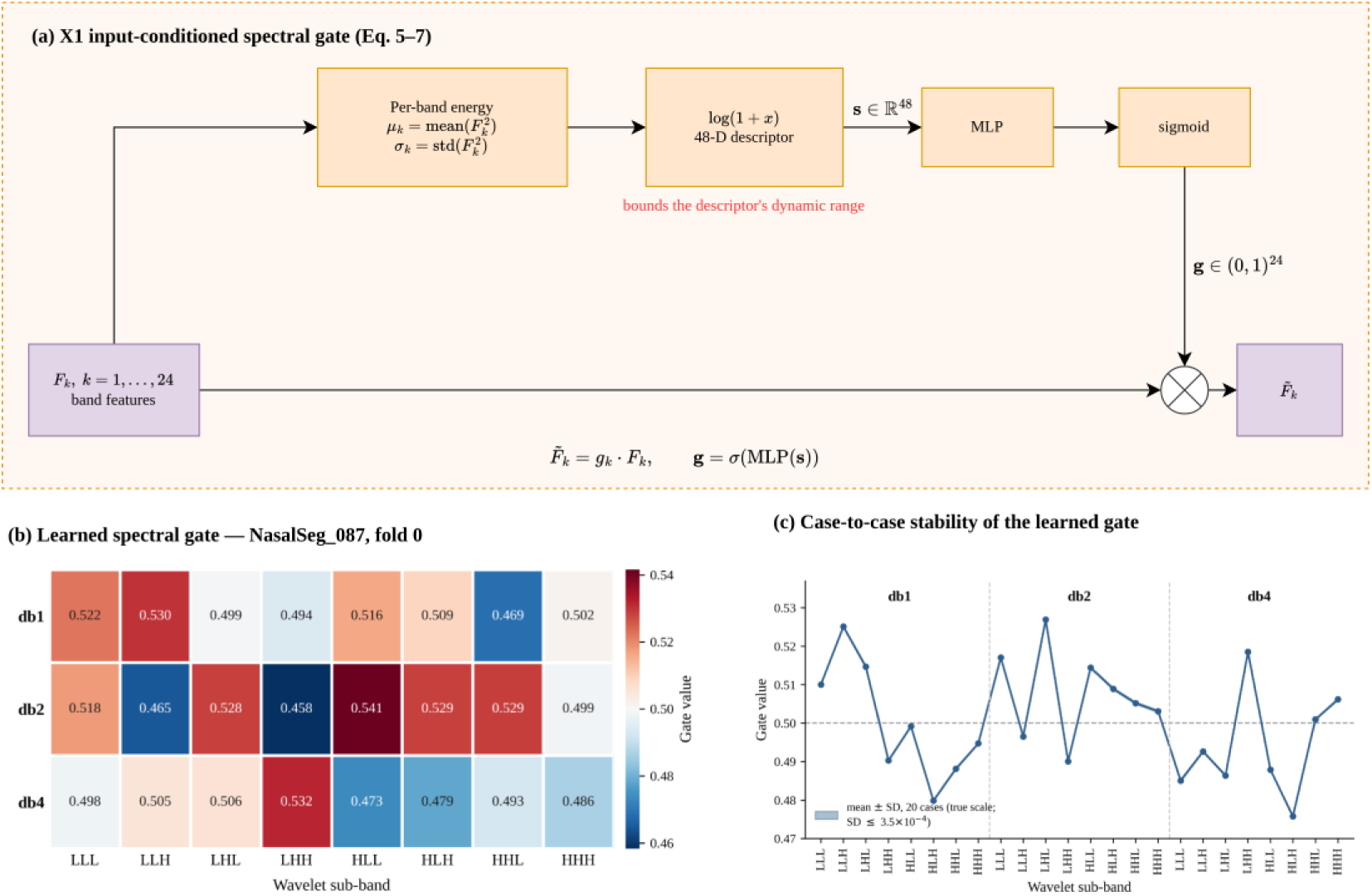
The X1 input-conditioned spectral gate. (a) Mechanism: the 24 band features F_k obtained from the wavelet sub-bands are summarised by their per-band energy (mean and standard deviation of F_k²), log-compressed and concatenated into a 48-dimensional descriptor s, and mapped by a small MLP with sigmoid output to 24 gate values g, which rescale the corresponding band features. The log(1 + x) step bounds the dynamic range of the descriptor before the MLP. (b) Gate values learned for a representative test case (NasalSeg_087, fold 0), arranged by wavelet basis and directional sub-band; colour is centred at 0.5, the value at which a band is neither boosted nor suppressed, and the observed range is 0.458–0.541. (c) Case-to-case stability: three-fold mean gate value per sub-band, with a ±1 SD band computed across the 20 test cases.

Formally, the per-band energy of each band feature is taken as its elementwise square; the mean and standard deviation of this energy are computed for each of the 24 bands, log-compressed by log(1 + x), and concatenated into a 48-dimensional descriptor (Eq. 5). The descriptor is passed through a small MLP with sigmoid output to obtain 24 gate values (Eq. 6), which rescale the corresponding band features (Eq. 7).

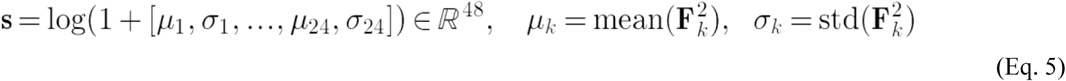

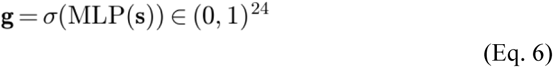

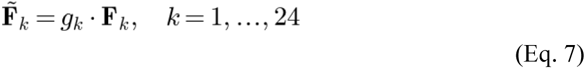

The objective variant WaveGate-X2 (abbreviated X2 below) leaves the WaveGate-Base architecture unchanged and instead adds a boundary loss [14], computed from the signed distance map of the ground-truth foreground and integrated against the predicted probability at the finest decoder resolution only, with its weight annealed linearly from 0 to 0.2 over the first 60 epochs. This is a strict addition to the baseline objective: nnU-Net’s deep supervision is retained unchanged, with the Dice + cross-entropy loss applied at every supervised decoder scale under the default 1/2^i weighting, and the boundary term is attached solely to the finest of these scales. The proposed model, WaveGate-nnU-Net (WaveGate), combines X1 and X2 with no additional change, testing whether their combination provides additional benefit beyond either component alone.

Formally, the boundary term integrates the predicted foreground probability against a signed distance map of the ground truth (Eq. 8), with its weight annealed linearly over the first 60 epochs (Eq. 9), giving the full X2 training objective (Eq. 10). WaveGate applies this objective to the X1-gated representation with no further change (Eq. 11).

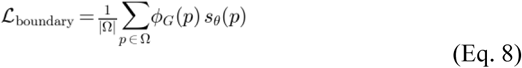

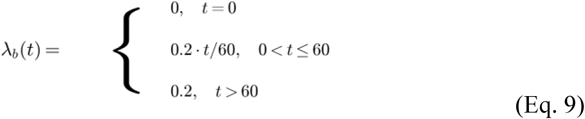

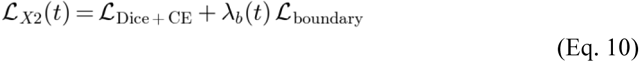

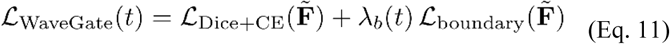

All models were implemented through a common Protocol/Adapter framework with standardized model interfaces and admission tests for tensor shape, gradient reachability, and tiny-sample overfitting; cross-architecture compatibility was additionally verified with U-Mamba [6].

### 2.3 Statistical validation and analysis

For the reduced-data experiment, Protocol B was constructed as a 3-fold partition of the 110 development cases, with mutually exclusive validation folds of 37/37/36 cases and corresponding training sets of 73/73/74 cases. Each development case served as validation exactly once, while the held-out 20-case test set remained unchanged and was excluded from split construction. This design reduced the number of training cases by approximately 19% relative to Protocol A while preserving complete validation coverage of the development set.

The WaveGate-versus-matched-baseline comparison under Protocol B was treated as the primary analysis; Protocol A arm contrasts and pairwise X1/X2/WaveGate comparisons were treated as secondary analyses. The primary reduced-data summary was the mean WaveGate-minus-baseline Dice difference across the three folds, reported with a fold-level 95% confidence interval and paired standardized effect size (Cohen’s d_z). Because the training sets overlap by construction in k-fold cross-validation, the conventional fold-level paired t-test was treated as descriptive and supplemented by a Nadeau-Bengio corrected analysis [15]. Interpretation therefore emphasized effect estimates, confidence intervals, and consistency across folds rather than a single p-value.

As a secondary case-level analysis, the 60 repeated test observations (20 cases x 3 folds) were analyzed using a mixed-effects model with arm as a fixed effect and case and fold as random intercepts. Because only three fold levels were available, a sensitivity analysis treating fold as fixed and using case-clustered standard errors was also performed. Directional consistency across individual test cases was additionally assessed using a sign test.

The model is specified as Eq. 12, with crossed random intercepts for test case and fold; β is the quantity of interest.

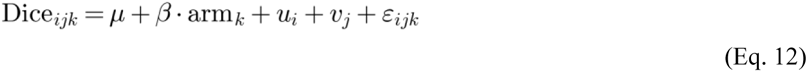

For Protocol A, case-level arm-versus-baseline comparisons were evaluated using paired tests and corrected for multiplicity across v2, X1, X2, and WaveGate using the Holm-Bonferroni procedure [16]. Equivalence between WaveGate and the individual X1 and X2 variants was evaluated using two one-sided tests with a ±0.10-percentage-point Dice equivalence margin [17], chosen as a conservative threshold for a practically negligible difference.

## 3. Results

### 3.1 Full-data protocol

Against the matched nnU-Net baseline (95.59% Dice), all corrected and investigational variants showed positive Dice differences: v2 +0.12 pp, X1 +0.17 pp, X2 +0.18 pp, and WaveGate +0.19 pp. WaveGate achieved the largest improvement (95.78% Dice; 95% CI [0.09, 0.29]; Holm-adjusted p = 0.004). Wilcoxon signed-rank testing also supported the improvements for v2, X2, and WaveGate, while X1 remained borderline (p = 0.064). Thus, the full-data protocol consistently favored the integrated variants, although the absolute gains were modest.

**Table 1.** Single-fold results on the held-out test set (n = 20), full 91/19/20 protocol (Protocol A). Dice values are mean ± population SD.

| Model | Dice (%) | $\Delta$ vs. baseline (pp) | 95% CI (pp) | d_z | p (Holm) | Wilcoxon p |
| --- | --- | --- | --- | --- | --- | --- |
| nnU-Net baseline | 95.59 ± 1.06 | — | — | — | — | — |
| WaveGate-Base (v2) | 95.71 ± 1.05 | +0.120 | [+0.029, +0.211] | 0.62 | 0.0253 | 0.0136 |
| WaveGate-X1 (spectral gate) | 95.76 ± 1.04 | +0.168 | [+0.020, +0.315] | 0.53 | 0.0279 | 0.0637 |
| WaveGate-X2 (boundary objective) | 95.77 ± 1.07 | +0.182 | [+0.072, +0.292] | 0.77 | 0.0079 | 0.0023 |
| WaveGate (proposed, X1 + X2) | 95.78 ± 1.07 | +0.190 | [+0.088, +0.292] | 0.87 | 0.0039 | 0.00085 |

After harmonizing the physical-space boundary evaluation across all arms, all three augmented variants also showed lower ASD than the baseline. WaveGate reduced ASD from 0.2516 to 0.2348 mm (−0.0169 mm, 95% CI [−0.0279, −0.0058]), corresponding to an approximately 6.7 % relative reduction in mean boundary error.

**Table 2.** Boundary metrics on the held-out test set (n = 20), full protocol, after harmonized physical-space evaluation. Δ ASD is arm minus baseline; negative values indicate lower boundary error. Δ ASD and 95% CIs were computed from unrounded per-case values prior to rounding for display; subtracting the rounded arm-level means shown in this table may therefore not reproduce Δ to the last decimal.

| Model | Dice (%) | ASD (mm) | $\Delta$ ASD (mm) | 95% CI (mm) | d_z | p (Holm) | Wilcoxon p |
| --- | --- | --- | --- | --- | --- | --- | --- |
| nnU-Net baseline | 95.59 ± 1.06 | 0.2516 | — | — | — | — | — |
| WaveGate-X1 | 95.76 ± 1.04 | 0.2364 | −0.0152 | [−0.0277, −0.0028] | 0.57 | 0.0191 | 0.0083 |
| WaveGate-X2 | 95.77 ± 1.07 | 0.2368 | −0.0148 | [−0.0253, −0.0043] | 0.66 | 0.0163 | 0.00026 |
| WaveGate | 95.78 ± 1.07 | 0.2348 | −0.0169 | [−0.0279, −0.0058] | 0.71 | 0.0145 | 0.000063 |

The component analysis showed that X1, X2, and WaveGate were pairwise equivalent within a ±0.10 pp Dice margin, indicating limited additional peak-accuracy benefit from combining spectral gating with the boundary objective.

### 3.2 Core result: robustness under reduced training data

The strongest effect emerged when the amount of training data was reduced. Under Protocol B, the nnU-Net baseline decreased from 95.59% Dice with 91 training cases to 92.88% with 73-74 cases, a loss of 2.71 pp. In contrast, WaveGate remained essentially unchanged, decreasing only from 95.78% to 95.75% (−0.03 pp). The resulting gap between the two models therefore expanded from only +0.19 pp under the full-data protocol to +2.88 pp under reduced training data.

Across the three reduced-data folds, WaveGate outperformed the matched baseline by +2.80, +2.08, and +3.75 pp, respectively. The cross-fold mean advantage was +2.88 pp (fold-level 95% CI [0.80, 4.96], n = 3 folds; Cohen’s d_z = 3.43), with the effect pointing in the same direction in all three folds. The consistency of this effect is particularly notable because the reduction in training data affected the baseline substantially while leaving WaveGate almost unchanged.

Several checks were performed to determine whether this pattern could be explained by training artifacts. The baseline validation curves showed no sustained late-epoch improvement, providing no evidence that the reduced-data baseline was simply undertrained. In addition, nnU-Net used the same 250 gradient steps per epoch regardless of training-set size, so the reduced-data comparison did not receive less optimization effort. Finally, all fold-specific models were verified as independently trained checkpoints, ruling out accidental reuse of the same model across folds.

Because the fold training sets overlap by construction, the reduced-data result was evaluated using several complementary statistical views rather than a single significance test. The descriptive fold-level paired t-test gave p = 0.027, while the dependence-aware Nadeau-Bengio correction gave p = 0.064. The magnitude of the effect, however, was highly stable when analyzed at the case level. A mixed-effects model over the 60 repeated case-fold observations estimated the same +2.88 pp advantage (95% CI [2.12, 3.64]), and a fold-fixed model with case-clustered standard errors again produced the same point estimate, with a 95% CI of [1.80, 3.96].

The direction of improvement was also remarkably consistent at the individual-case level. WaveGate exceeded the baseline in all 60 case-fold comparisons and, after averaging across the three folds, in all 20 held-out test cases (sign test, p = 1.9×10⁻⁶). Per-case Dice improvements ranged from +0.68 to +9.91 pp, and no test case showed a lower Dice score under WaveGate. This consistency indicates that the cross-fold mean advantage was not driven by only a small subset of difficult cases.

**Table 3.** True 3-fold cross-validation (Protocol B). Training sets contain 73–74 cases, approximately 19% fewer than the 91-case Protocol A. Test set n = 20, identical across folds.

| Fold | n_train | n_val | nnU-Net baseline Dice (%) | WaveGate Dice (%) | $\Delta$ (pp) |
| --- | --- | --- | --- | --- | --- |
| Fold 0 | 73 | 37 | $92.94 \pm 2.47$ | $95.74 \pm 1.10$ | +2.80 |
| Fold 1 | 73 | 37 | $93.72 \pm 1.49$ | $95.80 \pm 1.17$ | +2.08 |
| Fold 2 | 74 | 36 | $91.97 \pm 5.04$ | $95.72 \pm 1.16$ | +3.75 |
| Cross-fold mean | 73.3 | 36.7 | $92.88 \pm 0.88$ | $95.75 \pm 0.04$ | +2.88 |

Surface-distance analysis supported the same overall pattern. Across the 60 case-fold observations, ASD decreased from 0.420 mm for the baseline to 0.237 mm for WaveGate (Δ = −0.184 mm, 95% CI [−0.230, −0.137]), showing that the reduced-data advantage was accompanied by improved boundary localization rather than Dice overlap alone.

**Figure 4.**
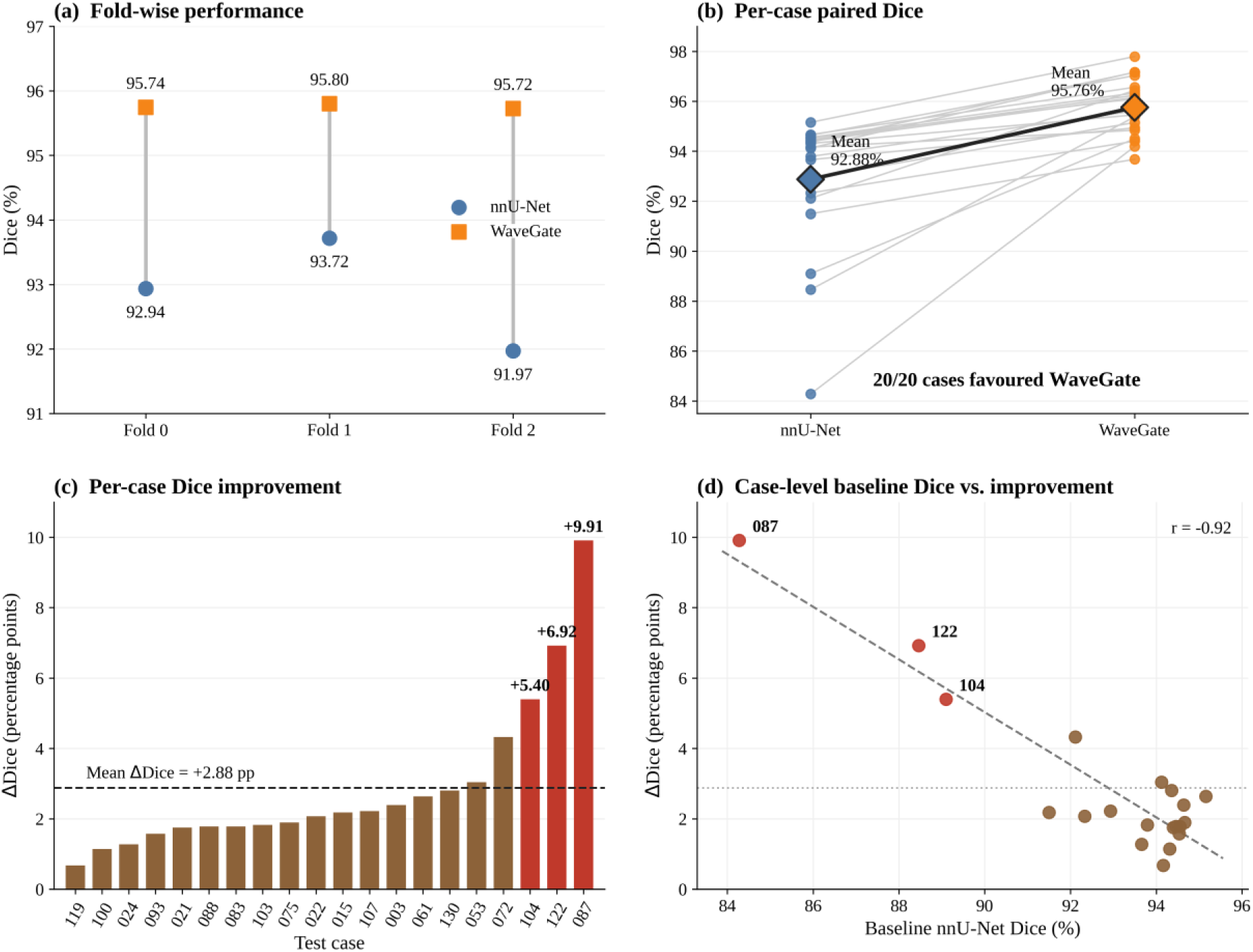
Performance of WaveGate under the reduced-data three-fold protocol. (a) Fold-wise Dice scores for the nnU-Net baseline and WaveGate across the three independently trained folds. (b) Paired case-level Dice scores averaged across the three fold-specific evaluations; WaveGate exceeded the baseline in all 20 test cases. (c) Case-wise Dice improvement of WaveGate over the baseline, ordered by magnitude (mean +2.88 pp; range +0.68 to +9.91 pp). (d) Exploratory case-level relationship between baseline Dice and the corresponding improvement; the dashed line is a descriptive trend only.

The models also differed substantially in their stability across folds. The baseline showed a cross-fold SD of 0.88%, while its within-fold SD varied widely from 1.49% to 5.04%, indicating substantial sensitivity to which cases were available for training. In contrast, WaveGate had a cross-fold SD of only 0.04%, and its within-fold SD remained tightly bounded between 1.10% and 1.17% across all three folds.

Taken together, the reduced-data experiment changes the interpretation of the method. Under the full-data protocol, WaveGate provides only a modest peak-accuracy improvement of +0.19 pp. When the training set is reduced by approximately 19%, however, the baseline loses 2.71 pp while WaveGate preserves nearly the same performance, producing a +2.88 pp advantage that is consistent across folds, cases, and boundary-error measurements. The main practical value of WaveGate therefore lies in maintaining segmentation accuracy and reducing sensitivity to training-set composition when fewer annotated CT volumes are available.

**Figure 5.**
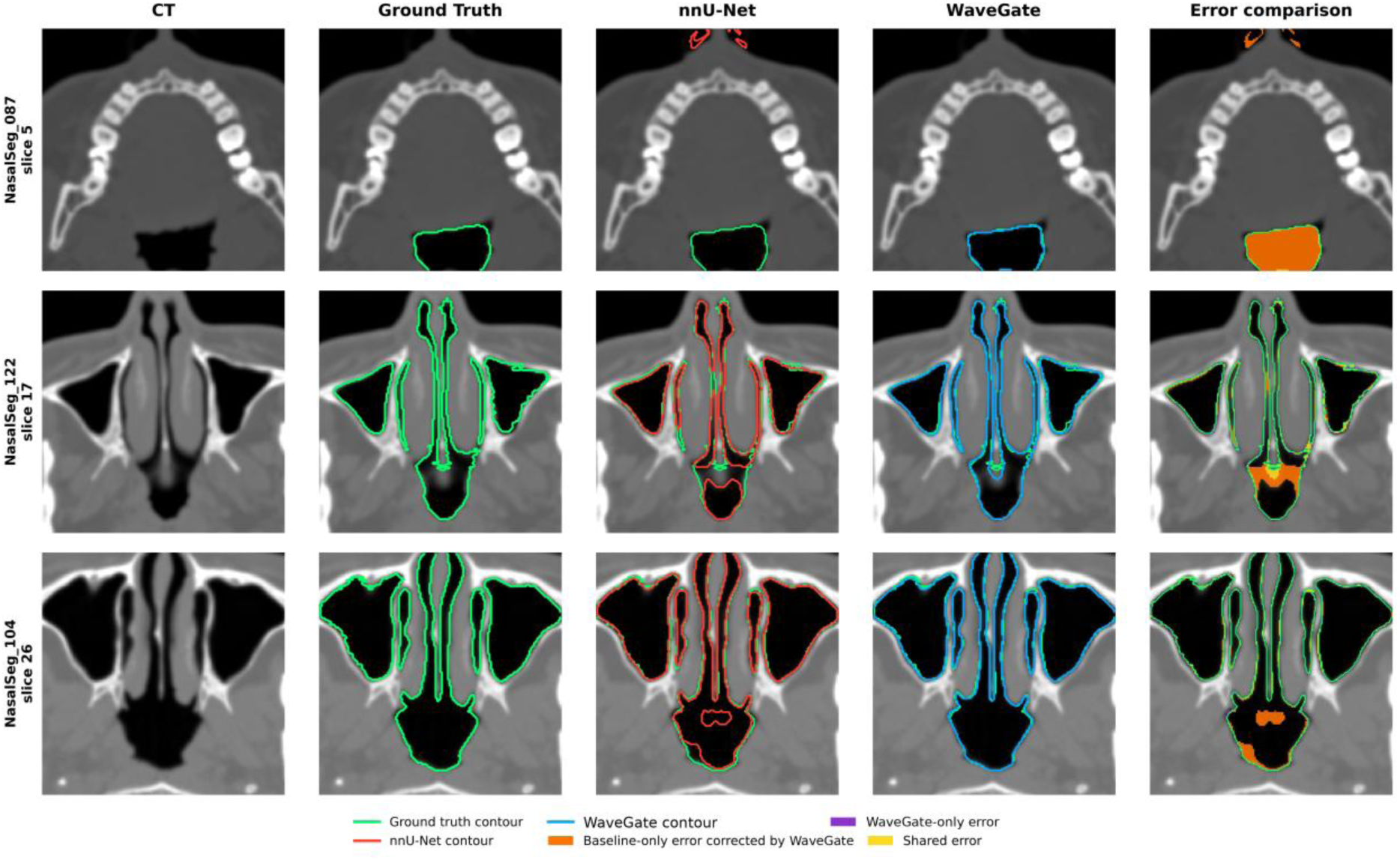
Qualitative comparison of three-fold consensus segmentations under the reduced-data protocol. Representative axial CT slices are shown for the three cases with the largest improvements. Ground-truth boundaries are shown in green, nnU-Net predictions in red, and WaveGate predictions in blue. Orange denotes baseline-only errors corrected by WaveGate; purple denotes errors introduced only by WaveGate; yellow denotes errors shared by both methods.

## 4. Discussion

### 4.1 Clinical relevance of data efficiency

Under full-data training, WaveGate improved Dice over the matched nnU-Net baseline by 0.19 pp. More importantly, its advantage became substantially larger when training data were reduced. With approximately 19% fewer training cases, WaveGate retained essentially the same performance as under the full-data protocol (95.78% vs. 95.75%), whereas the baseline declined from 95.59% to 92.88%. This produced a +2.88 pp advantage for WaveGate under reduced-data training, with WaveGate outperforming the baseline in all 20 test cases. Together, these findings indicate that the principal benefit of the proposed pipeline is not merely a small increase in peak segmentation accuracy, but substantially greater robustness to reduced training-data availability.

This property is particularly relevant to nasal-sinus CT segmentation, where volumetric annotation is demanding because of complex anatomy, thin bony boundaries, and narrow anatomical structures. Maintaining near-full-data performance with fewer annotated training cases therefore represents a practically meaningful form of annotation efficiency. The consistent improvement across all 20 test cases further suggests that the reduced-data advantage is not driven by a small subset of unusually difficult cases, but reflects a systematic benefit across the held-out test cohort. A similar data-dependent advantage has been observed for attention gating in abdominal CT segmentation, where the baseline deteriorated more strongly as training data were reduced [4]. Our results extend this observation to nasal-sinus CT and show that the proposed frequency-enhanced pipeline is particularly advantageous in the limited-data regime.

The ablation results further help interpret this behavior. Under full-data training, X1, X2, and WaveGate reached a similar performance range, suggesting that abundant training data allow nnU-Net to approach a common performance ceiling. When training data become more limited, however, the complete WaveGate pipeline preserves performance while the baseline deteriorates substantially. This pattern is consistent with the frequency-domain branch providing an additional structured representation that complements spatial learning when supervision is reduced. In particular, wavelet decomposition explicitly exposes information across multiple frequency scales, including high-frequency components relevant to thin boundaries and fine anatomical structures. The resulting multi-scale frequency representation may therefore act as an inductive prior that reduces the amount of training data required to learn these structures reliably. From this perspective, the +2.88 pp reduced-data advantage provides evidence that the practical contribution of WaveGate emerges most clearly under limited-data conditions, where its frequency-enhanced representation helps sustain segmentation performance.

### 4.2 Equivalence rather than synergy

X1 changes the architecture and X2 changes the objective, yet WaveGate is statistically equivalent to either alone within a margin narrower than each component’s own effect against the baseline — an established equivalence finding; it suggests that X1 may warrant consideration as a lower-cost alternative under full-data conditions, although its reduced-data robustness was not evaluated. WaveGate, not X1, was carried into the reduced-data validation because its full-data advantage was confirmed by both the t-test and the Wilcoxon test, whereas X1’s Wilcoxon result was borderline, despite X1 dominating on a cost basis.

### 4.3 Limitations

The 20-case test set was periodically inspected at developmental milestones (e.g., diagnosing the v1 gating deadlock), which is weaker than direct hyperparameter tuning but may still introduce mild optimistic bias, particularly into the small full-data margins. Although Protocols A and B draw from the same 110-case development cohort and use the same 20-case held-out test set, each model is trained on a different number of cases (91 versus 73-74); cross-protocol changes are therefore interpreted descriptively rather than as paired inferential comparisons. Our reconstruction of the frequency-domain components of [12] differs from the published implementation in band-weight normalization, attention scaling, router input composition, and wavelet differentiability, and its parameter count is closer to the lightweight published variant than to the 414.57M-parameter full variant; statements about the frequency-domain mechanism should therefore be read as statements about this reconstruction rather than about the published network. Fold-level inference rests on three non-independent, pairwise-overlapping folds, for which the Nadeau-Bengio correction is the more conservative estimate (p = 0.064); we rest the conclusion on effect size, confidence intervals, and case-level consistency rather than the fold-level p-value alone, and identify a higher fold count as the appropriate remedy for future work. Finally, all results derive from a single dataset, and the reduced-data experiment subsamples the same development cohort; it therefore demonstrates within-distribution annotation efficiency, not generalization to an independently collected cohort, scanner population, or annotation protocol.

## 5. Conclusion

We asked whether frequency-domain modeling can be incorporated into nnU-Net in a lightweight form, and whether the resulting gains survive rigorous validation. Under the full protocol, three controlled variants — a spectral gate (X1), a boundary objective (X2), and their combination (WaveGate) — all showed small positive Dice differences relative to the matched baseline, with WaveGate achieving the largest gain (+0.19 pp; Holm-adjusted p = 0.004). Under the reduced-data three-fold protocol, the picture changed qualitatively: the baseline lost 2.71 pp while WaveGate lost only 0.03 pp, a +2.88 pp cross-fold advantage corroborated by surface distance and consistent across all 20 test cases. We therefore report within-distribution data efficiency of the combined pipeline, rather than peak accuracy or isolated frequency-mechanism efficacy, as the substantive contribution of this work.

## Acknowledgments

The authors gratefully acknowledge the creators of the NasalSeg dataset for providing open-access nasal cavity and paranasal sinus CT data that enabled the experimental evaluation in this study.

## Ethical approval

This study used the publicly available, de-identified NasalSeg dataset [1]. The original data collection and de-identification were approved by the Institutional Review Board of Huashan Hospital, Fudan University (No. KY2024-1345), which waived the requirement for informed consent given the retrospective, anonymized nature of the data [1]. As this study performs only secondary analysis of this publicly released, de-identified dataset and collected no new patient data, no additional institutional ethical approval was required.

## Data availability

The NasalSeg dataset analyzed in this study is publicly available via Zenodo (https://doi.org/10.5281/zenodo.13893419) and is described in Zhang et al. [1]; the original code repository is available at https://github.com/YichiZhang98/NasalSeg. The rebuilt 3-fold split, trained model checkpoints, and evaluation code produced in this study are available from the corresponding author upon reasonable request.

## CRediT authorship contribution statement

Yu-Chen Chen: Conceptualization, Methodology, Software, Formal analysis, Investigation, Validation, Visualization, Writing – original draft. Shu-Yen Wan: Conceptualization, Methodology, Supervision, Validation, Writing – review & editing, Funding acquisition.

